# Effects of household concrete floors on maternal and child health – the CRADLE trial: a randomised controlled trial protocol

**DOI:** 10.1101/2024.07.26.24311076

**Authors:** Mahbubur Rahman, Farjana Jahan, Suhi Hanif, Afsana Yeamin, Abul Kasham Shoab, Jason R. Andrews, Ying Lu, Sarah Billington, Nils Pilotte, Ireen S. Shanta, Mohammad Jubair, Mustafizur Rahman, Mamun Kabir, Rashidul Haque, Fahmida Tofail, Sakib Hossain, Zahid H Mahmud, Ayse Ercumen, Jade Benjamin-Chung

**Affiliations:** Environmental Health and WASH, International Centre for Diarrhoeal Disease Research, Bangladesh (icddr,b),Dhaka-1212, Bangladesh; Global Health and Migration Unit, Department of Women’s and Children’s Health, Uppsala University, Sweden; King Center on Global Development, Stanford University; Department of Epidemiology and Population Health, Stanford University and Chan Zuckerberg Biohub Investigator.; Division of Infectious Diseases and Geographic Medicine, Stanford University; Department of Biomedical Data Science, Stanford University; Department of Civil and Environmental Engineering, Stanford University; Department of Biological Sciences, Quinnipiac University; Genome Centre Infectious Diseases Division, International Centre for Diarrhoeal Disease Research, Bangladesh; Infectious Diseases Division, International Centre for Diarrhoeal Disease Research, Bangladesh; Maternal and Child Nutrition, International Centre for Diarrhoeal Disease Research, Bangladesh; Laboratory of Environmental Health, International Centre for Diarrhoeal Disease Research, Bangladesh; College of Natural Resources, North Carolina State University; Chan Zuckerberg Biohub, San Francisco

## Abstract

**Introduction:** Early life soil-transmitted helminth infection and diarrhea are associated with growth faltering, anemia, impaired child development, and mortality. Exposure to fecally contaminated soil inside the home may be a key contributor to enteric infections, and a large fraction of rural homes in low-income countries have soil floors. The objective of this study is to measure the effect of installing concrete floors in homes with soil floors on child soil-transmitted helminth infection and other maternal and child health outcomes in rural Bangladesh.

**Methods and analysis:** The Cement-based flooRs AnD chiLd hEalth (CRADLE) trial is an individually randomised trial in Sirajganj and Tangail districts, Bangladesh. Households with a pregnant woman, a soil floor, walls that are not made of mud will be eligible, and no plan to relocate for 3 years. We will randomise 800 households to intervention or control (1:1) within geographic blocks of 10 households to account for strong geographic clustering of enteric infection. Laboratory staff and data analysts will be blinded; participants will be unblinded. We will install concrete floors when the birth cohort is in utero and measure outcomes at child ages 3, 6, 12, 18, and 24 months.

The primary outcome is prevalence of any soil-transmitted helminth infection (*Ascaris lumbricoides*, *Necator americanus*, or *Trichuris trichiura*) detected by qPCR at 6, 12, 18, or 24 months follow-up in the birth cohort. Secondary outcomes include household floor and child hand contamination with *E. coli*, extended-spectrum beta-lactamase producing *E. coli*, and soil-transmitted helminth DNA; child diarrhea, growth, and cognitive development; and maternal stress and depression.

**Ethics and dissemination:** Study protocols have been approved by institutional review boards at Stanford University and the International Centre for Diarrheal Disease Research, Bangladesh (icddr,b). We will report findings on ClinicalTrials.gov, in peer-reviewed publications, and in stakeholder workshops in Bangladesh.

**Trial registration number:** NCT05372068, pre-results

**Strengths and limitations of this study:**

- Using a randomised design in a large sample will allow us to minimize potential confounding by household wealth, which may have influenced prior observational studies’ findings on concrete floors and health.
- Measurement of a diverse set of health outcomes within different domains (infections, antimicrobial resistance, child growth, cognitive development, mental health, quality of life) will capture broad potential benefits of the intervention.
- Longitudinal measurements will capture any variation in intervention impact as children learn to sit, crawl, walk and spend more time outdoors and their exposures change.
- Rich data on intermediate variables on household contamination and maternal bandwidth, time use, and mental health will allow us to investigate whether concrete floors influence child health and development primarily through environmental or maternal pathways.
- It is possible that child exposures outside the home will attenuate the effect of concrete floors on child health outcomes.

## Introduction

The United Nations has enshrined basic housing as a human right, and housing improvements are associated with improved health (Thomson et al. 2009; Thomson, Sellstrom, and Petticrew 2013). Yet, 58.8% of homes in low-income countries are unimproved, with soil floors and walls (Morakinyo, Fagbamigbe, and Adebowale 2022) made of palm or thatch, and few studies have investigated health benefits of housing upgrades in low- or middle-income countries (LMICs). In low-income homes, inadequate sanitation infrastructure can result in contamination of household soil and surfaces with human and animal feces. In particular, soil floors are a reservoir for enteric pathogens such as soil-transmitted helminths (STH), *Shigella*, and pathogenic *E. coli (Steinbaum et al. 2019, 2016; Shrivastava et al. 2020)*. Multiple studies have found that the amount of *E. coli* and STH is similar or higher on soil floors inside the home than on latrine floors (Steinbaum et al. 2016; Pickering et al. 2012; Schulz and Kroeger 1992). *Ascaris lumbricoides* eggs are extremely hardy and can survive for years in soil; other STH species can survive for weeks or months (S. Brooker, Clements, and Bundy 2006; Ransom and Foster 1920). Soil is also a reservoir of antimicrobial resistant pathogens because microorganisms in soil can exchange genes with those in feces deposited on soil (Zhu et al. 2019). Children frequently play, eat, and drink on floors that are contaminated with human or animal feces, and young children frequently ingest soil, increasing their risk of infection with STH, other enteric pathogens, and antimicrobial resistant pathogens (Kwong, Ercumen, Pickering, Arsenault, et al. 2020; Kwong, Ercumen, Pickering, Unicomb, et al. 2020; Goddard et al. 2020; Ercumen, Pickering, et al. 2018; Ercumen, Mertens, et al. 2018; Fuhrmeister et al. 2020; Pickering, Swarthout, et al. 2019; Kwong et al. 2021).

Collectively, enteric infections, including STH and diarrhea, are a major contributor to the global burden of disease. They were the third leading cause of death for children <5 years in 2019 (“Global Burden of Disease” 2019). Chronic STH infection is associated with an increased risk of child wasting, anemia, dysentery, intestinal obstruction, impaired child development, and mortality (Simon Brooker, Bethony, and Hotez 2004; Jourdan et al. 2018). Hookworm infection alone is responsible for up to $139 billion in annual productivity losses globally (Bartsch et al. 2016). In addition to increased risk of mortality, early life diarrhea is associated with growth faltering, anemia, and impaired child development (Pinkerton et al. 2016; Lorntz et al. 2006; Niehaus et al. 2002; Kotloff et al. 2013; MacIntyre et al. 2014; MAL-ED Network Investigators 2017; Kosek and MAL-ED Network Investigators 2017).

Existing interventions to reduce enteric infections, such as water, sanitation, and hygiene (WASH), do not address soil exposure, and a recent meta-analysis found that low-cost household WASH interventions did not reduce enteric pathogen prevalence in household soil samples (Mertens et al. 2023). Further, WASH interventions require sustained behavior-change promotion, which limits scalability (Pickering, Null, et al. 2019). In addition, recent WASH trials had only modest effects on diarrhea and mixed effects on STH infections (Pickering, Null, et al. 2019; Vaz Nery et al. 2019). Alternative interventions that reduce environmental pathogen reservoirs and do not require sustained behavior change are urgently needed to improve child health in LMICs.

Installing finished floors, such as concrete, in homes with soil floors is a promising potential intervention to reduce child enteric infection. Concrete floors are easier to keep clean than soil floors, which may contribute to lower levels of fecal pathogens on surfaces, and subsequently on hands and fomites. Concrete may also interrupt the life cycle of STH, which require soil to reach their infective stage (S. Brooker, Clements, and Bundy 2006). A recent meta-analysis of observational studies estimated that the odds of any enteric or parasitic infection was 0.75 times lower and the odds of helminth infections was 0.68 times lower in homes with improved floors (e.g., concrete, wood) vs. unimproved floors (e.g., soil) (Legge, Pullan, and Sartorius 2023). In addition, studies have found protective associations between improved floors and diarrhea (Koyuncu et al. 2020; Cattaneo et al. 2009).

By making homes more comfortable and easier to keep clean, floor upgrades can also improve quality of life and reduce stress. A study in Mexico found that installing concrete floors was associated with 10.6% lower stress, 12.5% lower depression, and 18.7% higher satisfaction with quality of life among mothers (Cattaneo et al. 2009). Slum upgrades, including concrete or wood floor installation, have been found to improve sleep, quality of life, and happiness (Galiani et al. 2017; Simonelli et al. 2013). Though findings from these studies are promising, all prior research has been observational, and associations may be confounded by household wealth.

Concrete floors could also influence child development through either an environmental pathway or maternal pathway (Figure 1). In the environmental pathway, installing concrete floors removes the majority of soil from the home’s interior and makes the home easier to clean, reducing child exposure to enteric pathogens, likely also reducing enteric infections, which are associated with impaired child development (Guernier et al. 2017; Grenham et al. 2011; Scharf, Deboer, and Guerrant 2014). In the maternal pathway, if concrete floors make the living environment more comfortable and easier to clean, this may result in increased maternal bandwidth (Mullainathan and Shafir 2013) – the ability to problem solve, recall information, reason logically, plan and allocate attention and initiate and control actions (Schilbach, Schofield, and Mullainathan 2016). Higher levels of maternal working and short-term memory are independently associated with maternal scaffolding behaviors when interacting with young children (Obradović et al. 2017, 2016). These scaffolding behaviors are associated with child cognitive skills at age four years (Obradović et al. 2016). Further, by reducing the time required to clean floors, concrete floors may increase mothers’ discretionary time. This may increase their ability to seek medical care and/or devote time to preventive health behaviors for themselves or their children, and engage in scaffolding behaviors that promote child development (Hyde, Greene, and Darmstadt 2020).

**Figure 1.**
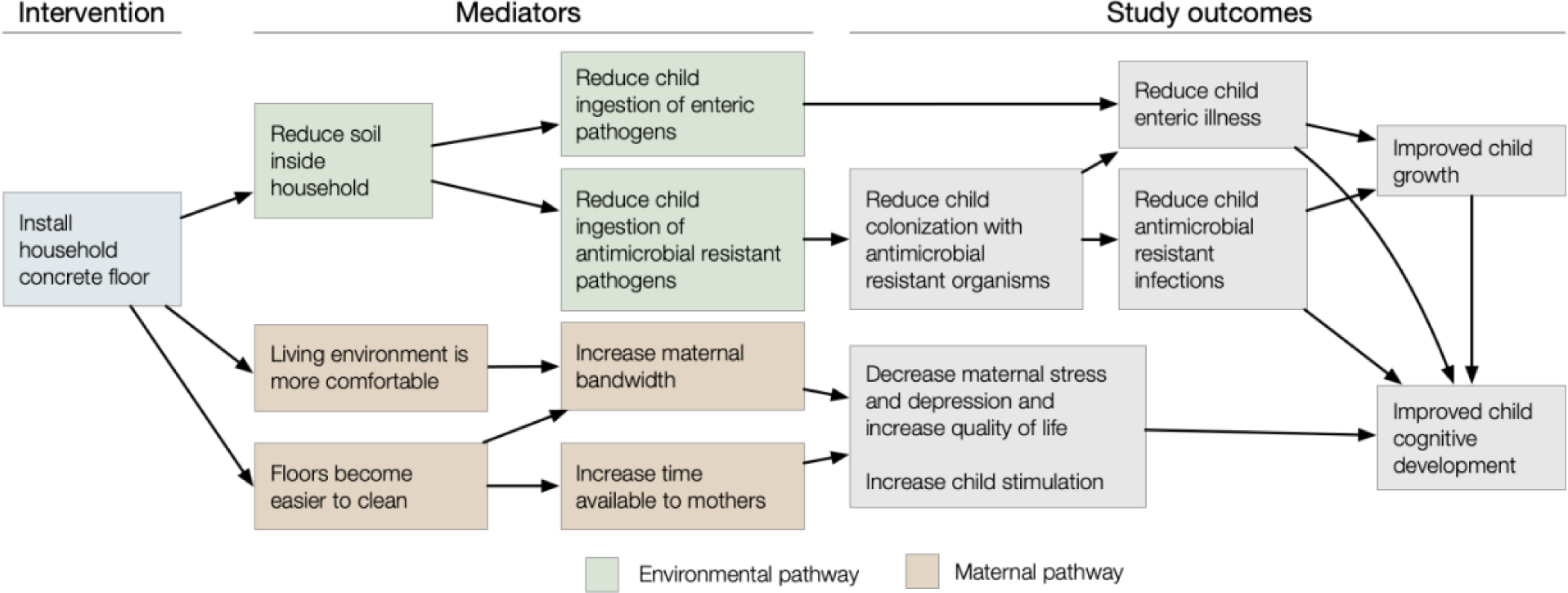
Hypothesized mechanisms of intervention effects.

### Objectives

Here, we describe the protocol for the Cement-based flooRs AnD chiLd hEalth (CRADLE) trial, a randomised trial in Sirajganj district, Bangladesh to determine whether concrete floors reduce child STH infection and diarrhea. Using a randomised design will minimize confounding that may have influenced prior observational studies. We will investigate the mechanism through which concrete floors influence health using rich environmental assessments, video observations of child activities, and biopsychosocial measurements of mothers. Our longitudinal design will allow us to investigate sustainability over 2 years and capture any variation in intervention impact as children learn to sit, crawl, walk and spend more time outdoors and their exposures change. Cost-effectiveness analyses incorporating maternal and child outcomes will inform decisions about whether to scale up concrete floor installation. This study will generate rigorous evidence to inform policies about whether concrete floors should be delivered as a health intervention in rural, low-income settings.

## Methods and analysis

### Study design

The CRADLE trial is an individually randomised trial in Sirajganj and Tangail districts, Bangladesh. The trial will enroll households with soil floors and a pregnant woman in her second or third trimester. For each block of 10 geographically contiguous households, we will randomise households 1:1 to intervention or control. We will install concrete floors when the birth cohort is in utero so that children in the intervention arm receive the intervention from birth. We will measure outcomes longitudinally when children are aged 0, 3, 6, 12, 18, and 24 months (Figure 2). While STH prevalence is generally higher among primary school-aged children, we chose to focus on the first two years of life, when children’s exposure to household floors is greatest. Between 6 and 24 months, children frequently sit, play, crawl, and eat on household floors and generally spend more time inside vs. outside the home. After this age, children’s outdoor exposures increase; while household concrete floors may still benefit older children, we expect that potential effects would be smaller. Additionally, the first 1000 days of life are considered critical for child development and therefore present an important window for health interventions (Victora et al. 2021).

**Figure 2.**
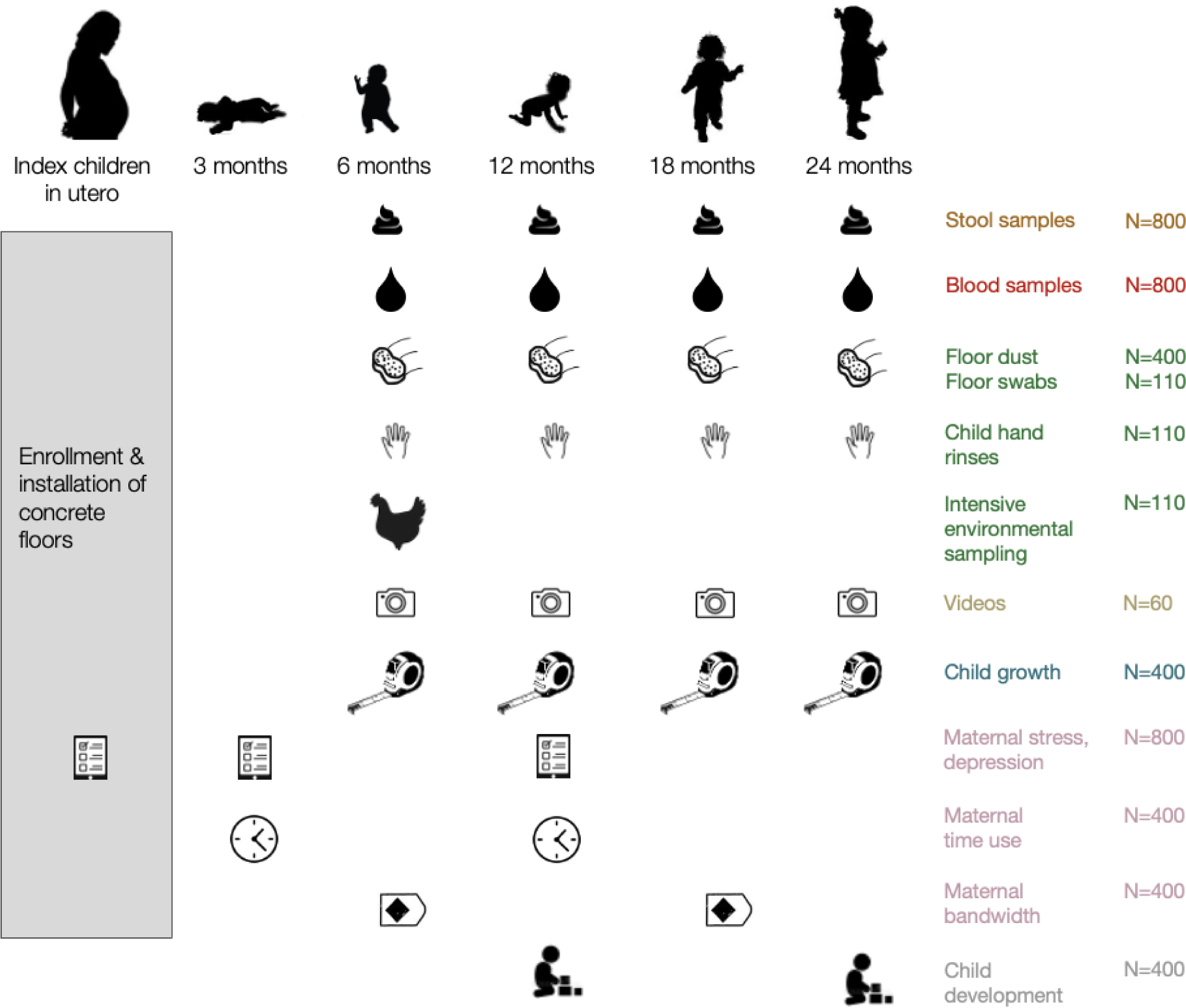
Planned measurements in the CRADLE trial. Intensive environmental samples collected at the 6-month follow-up include courtyard soil, floor soil, food, drinking water, chicken feces, cow feces.

### Study site

Our study site is located in Chauhali and Belkuchi upazilas (sub-districts) in Sirajganj district and Nagarpur upazila in Tangail district, Bangladesh (Figure 3). Chauhali upazila is approximately 210 km^2^ and includes 34,449 households, Belkuchi upazila is approximately 164 km^2^ and includes 96,110 households, and Nagarpur upazila is approximately 267 km^2^ and includes approximately 83,885 households (“Population and Housing Census 2022, National Report” 2023, “Bangladesh National Portal,” n.d.). Research staff from the International Centre for Diarrhoeal Disease Research, Bangladesh (icddr,b) will enroll households in all unions of Chauhali and Belkuchi and unions adjacent to Chauhali in Nagapur. Approximately 66% of households in our study area have mud floors, and the region is vulnerable to flooding and erosion (“Nationwide Climate Vulnerability Assessment in Bangladesh” 2018; Ali et al. 2017). According to the Chauhali Upazila Health & Family Planning Office, there is a high percentage of open defecation in this region, and clinical diagnoses of soil-transmitted helminth (STH) infections are common. The primary STH control strategy in Bangladesh consists of a biannual school-based deworming campaign whereby a single dose of mebendazole is administered to school-aged children (Hafiz et al. 2015; Gerber et al. 2023).

**Figure 3.**
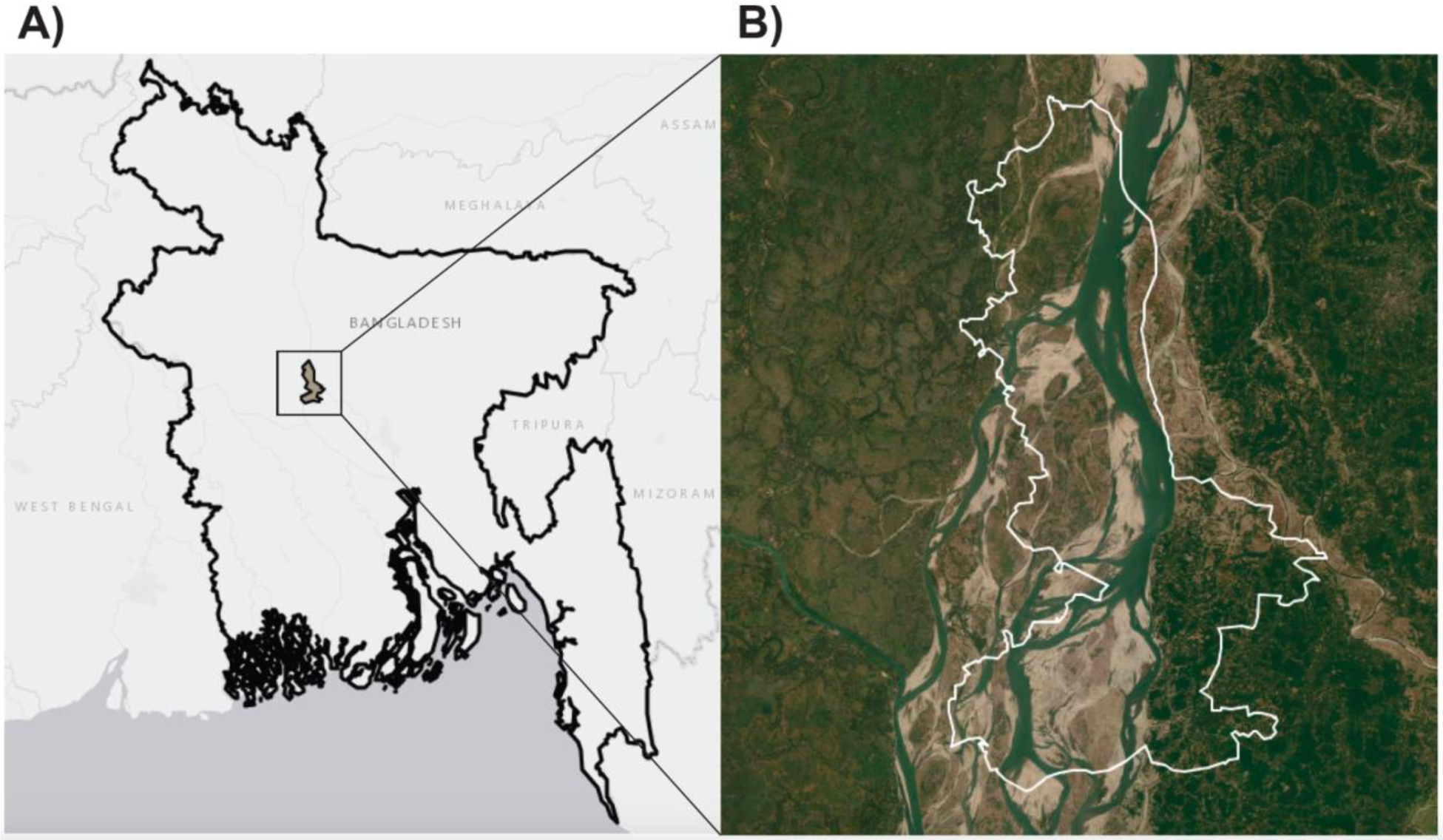
CRADLE Study site map. A) Map of study site location within Sirajganj and Tangail districts, Bangladesh. B) Exterior boundary of unions included in the study site in Chauhali, Belkuchi, and Nagarpur unions of Sirajganj and Tangail districts. Base map source: Esri.

### Inclusion and exclusion criteria

We will enroll households if they have floors made entirely of soil or earth and if there is a pregnant woman at 14-30 weeks of gestation at the time of enrollment. Pregnancy status will be self-reported. Primary outcomes will be measured in the child (or children, in the case of multiples) born to enrolled women (i.e., “index children”). We will exclude households with plans to relocate within 3 years or pregnant women who will be leaving their homes for child delivery for more than three months. Additionally, we will exclude households with mud walls as installing concrete floors may reduce structural stability.

### Outcomes

Our primary outcome is the prevalence of any of the following STH infections, *Ascaris lumbricoides, Necator americanus, or Trichuris trichiura,* detected using qPCR in child stool at 6, 12, 18, or 24 months after the birth of the index child. Secondary outcomes include the prevalence of each STH species by qPCR at child ages 6, 12, 18, or 24 months and caregiver-reported diarrhea in the past 7 days and diarrhea confirmed using stool consistency (Lane et al. 2011) at each follow-up. Additional secondary outcomes are listed in Table 1.

**Table 1.** Outcome measures.

|  |  |
| --- | --- |
| **Primary outcome** |  |
| Prevalence of any soil-transmitted helminth using qPCR | Proportion of children in which *Ascaris lumbricoides*, *Necator americanus*, or *Trichuris trichiura* was detected in stool by qPCR at any follow-up measurement (6, 12, 18, or 24 months of age). |
| **Secondary outcomes** |  |
| *Child health outcomes* |  |
| Prevalence of *Ascaris lumbricoides*, *Necator americanus* or *Trichuris trichiura* using qPCR | Proportion of children in which *Ascaris lumbricoides*, *Necator americanus* or *Trichuris trichiura* was detected in stool by qPCR at any follow-up measurement (6, 12, 18, or 24 months of age). |
| Mean *Ascaris lumbricoides*, *Necator americanus* and *Trichuris trichiura* eggs per gram | Geometric and arithmetic mean of *Ascaris lumbricoides*, *Necator americanus* and *Trichuris trichiura* eggs per gram detected in stool by Kato-Katz at any follow-up measurement (6, 12, 18, or 24 months of age) |
| Diarrhea prevalence | Prevalence of caregiver-reported and stool consistency based child diarrhea in the past 7 days (at least three loose or watery stools within 24 hours or at least one bloody stool) at any follow-up measurement (6, 12, 18, or 24 months of age) |
| Relative abundance and diversity of human pathogens | Human pathogens detected in child stool using shotgun metagenomic sequencing each follow-up round |
| Relative abundance and diversity of antimicrobial resistance genes | Antimicrobial resistance genes detected in child stool using shotgun metagenomic sequencing each follow-up round |
| *Child behavior* |  |
| Rate of child soil contact and ingestion | Mean number of soil contact and ingestion events per child-hour observed during 6-hour video observations at each follow-up (6, 12, 18, 24 months) |
| *Child development outcomes* |  |
| Mean ASQ-i score at 12 and 24 months | Mean communication, fine motor, gross motor, problem solving, personal-social and combined scores measured using the Ages and Stages Inventory calculated separately at 12 months and 24 months |
| Mean overall Family Care Indicator score at 12 and 24 months | Mean Family Care Indicator score overall measured separately at 12 months and 24 months (range: 0-3) |
| *Maternal outcomes* |  |
| Prevalence of stress (mild, moderate and severe) | Proportion of mothers with Perceived Stress Scale scores 0-13 (mild), 14-26 (moderate), 27-40 (severe) at 6 and 18 months |
| Prevalence of prenatal and postnatal depression | Proportion of mothers with Edinburgh Postnatal Depression scale score of 9.5 or higher at baseline and at 6 and 18 months |
| Prevalence of maternal satisfaction with quality of life, household floor quality and housing quality | Proportion of mothers reporting that they were “very satisfied”, “satisfied”, or “somewhat satisfied” with their quality of life, household floor quality and housing quality at each follow-up (6, 12, 18, 24 months) |
| Prevalence of improved maternal satisfaction with quality of life, household floor quality and housing quality | Proportion of mothers with an improvement in their reported quality of life, household floor quality and housing quality between baseline and each follow-up (6, 12, 18, 24 months) |
| Mean memory span | Mean length of consecutive blocks mothers correctly recalled (Corsi Span) using the Corsi Block Span task between baseline and follow-up at 3 and 12 months |
| Mean non-verbal reasoning score | Mean maternal Raven's Progressive Matrices score between baseline and follow-up at 3 and 12 months |
| Mean maternal cognitive inhibition | Mean percentage of correct responses and response time in the Hearts and Flowers task at 6 and 18 months |
| Mean maternal cognitive flexibility and attention shifting | Mean percentage of correct responses and response time in the Dimensional Change Card Sort test at 7 and 18 months |
| Mean maternal daily discretionary time | Maternal time poverty using hybrid time diaries and activity lists that measure discretionary time use at 3 and 12 months |
| *Environmental outcomes* |  |
| Prevalence of *Ascaris lumbricoides*, *Necator americanus*, and *Trichuris trichiura* in household floors | *Ascaris lumbricoides*, *Necator americanus*, and *Trichuris trichiura* detected by qPCR in household floor dust at any follow-up measurement (6, 12, 18, 24 months) |
| Prevalence of larvated *Ascaris lumbricoides* and *Trichuris trichiura* ova in household floors | Larvated *Ascaris lumbricoides* and *Trichuris trichiura* detected by microscopy in household floor swabs at any follow-up measurement (6, 12, 18, 24 months) |
| Most probable number of *E. coli* in household floor samples and child hand rinses | *E. coli* enumerated by IDEXX in household floor swabs and child hand rinses at any follow-up measurement (6, 12, 18, 24 months) |
| Most probable number of cefotaxime-resistant *E. coli* in household floor samples | Cefotaxime-resistant *E. coli* enumerated by IDEXX with cefotaxime supplement in household floor swabs at any follow-up measurement (6, 12, 18, 24 months) |
| Prevalence of extended-spectrum beta-lactamase producing *E. coli* (ESBL-E) and ESBL coding genes in household floor samples, courtyard soil, child hand rinses, drinking water, prepared food, cow feces and chicken feces | ESBL-E and ESBL coding genes in floor swabs, courtyard soil, child hand rinses, drinking water, prepared food, cow feces and chicken feces at 6 months follow-up |
| Mass of floor dust (g per m<sup>2</sup>) | Mass of dust on floors collected by sweeping a 2 m<sup>2</sup> floor area next to bed where child sleeps at night (6, 12, 18, 24 months) |
| Relative abundance and diversity of human pathogens | Human pathogens detected in floor soil, cow feces and chicken feces samples using shotgun metagenomic sequencing each follow-up round |
| Relative abundance and diversity of antimicrobial resistance genes | Antimicrobial resistance genes detected in floor soil, cow feces and chicken feces samples using shotgun metagenomic sequencing each follow-up round |
| *Floor outcomes* |  |
| Rate of floor sweeping and refinishing | Mean number of times floors were swept per household-day at each follow-up measurement and mean number of times floors were refinished per household-month at each follow-up measurement (6, 12, 18, 24 months) |
| Prevalence of holes or cracks in household floors | Proportion of household floors with holes or cracks at each follow-up measurement (6, 12, 18, 24 months) |
| Prevalence of impermeable household floors | Proportion of household floors in which surface water was still present after 1 minute at each follow-up measurement (6, 12, 18, 24 months) |

### Sample size calculation

The overall trial sample size was determined based on the primary outcome of prevalence of any STH at any follow-up (ages 6, 12, 18, 24 months). We estimated the required sample size using the standard formula for comparisons of two groups with repeated measures of a binary outcome (Liu and Liang 1997). From 2017-2020 in Sirajganj District, prevalence of *Ascaris lumbricoides* was 25.1%, and prevalence of *Trichuris trichiura* was <10% (Gerber et al. 2023). To be conservative, we assumed the prevalence of any STH in the control group over four follow-up rounds would be 20%. In our prior analyses of the association between concrete floors and any STH, the unadjusted prevalence ratio was 0.40 (95% CI 0.28, 0.56) (Benjamin-Chung et al. 2021), and the adjusted prevalence ratio was 0.73 (95% CI 0.52, 1.01) (Benjamin-Chung et al. 2015). We assumed a minimum detectable prevalence ratio of 0.70. We assumed intraclass correlation = 0.066 for repeated measurements of STH prevalence among children < 2 years (Platts-Mills et al. 2015). Assuming a Type I error = 0.05 and Type II error = 0.20, for a two-sided hypothesis test, the required number of households per arm is 336. Inflating this to account for 15% loss to follow-up and 10 households per randomisation block, the total required sample size across both arms is 800. 15% loss to follow-up was assumed based on the findings from the prior trial carried out by our team in rural Bangladesh (Luby et al. 2018), in which 6.5% of pregnant women miscarried or had still births, 3.5% of children died by 12-month follow-up, and <1% died between 12-24 months follow-up.

### Patient and public involvement

To support the translation of the trial’s findings into programs and policies, we will conduct stakeholder engagement workshops in Bangladesh during the trial preparation phase and results dissemination. We will engage different stakeholders from health, housing and construction sectors (Government of Bangladesh, engineering institutes, NGOs in housing and shelters, commercial companies on sustainable construction materials) and stakeholders working in climate change climate adaptation strategies for housing, shelters and water and sanitation. During the first workshop, we will gain input on proposed concrete floor design, health outcome measurement, and planned research outputs from stakeholders. We will also identify the gaps and areas for improvement in current policies, programs, and resources related to housing in rural Bangladesh. In both workshops, we will discuss bottlenecks and potential barriers to scaling up a potential improved flooring intervention and challenges related to sourcing sustainable materials for household floors.

The study team will convene community engagement meetings in each union and upazila in the study site. Attendees will include administrative officials, elected members, religious leaders, and local non-elected leaders. To build trust with the community, the study team will introduce study objectives, discuss potential community benefits, explain the randomisation process and intervention implementation plan, and address any concerns. If community leaders agree, then the team will proceed with recruitment. icddr,b will meet with leaders every 6 months to provide updates on study activities and learn about any other ongoing interventions in the study area.

### Enrollment

Field staff will select a random location in each union using spatial software. If a union spans both sides of the Jamuna river or one of its distributaries, a new random location will be used on each side of the river. The eligible household nearest to the random location will be selected as the first household. Field staff will discuss the trial with adults in the eligible household, including pregnant mothers, and obtain written informed consent (Appendix 1). Field staff will then spin a bottle and move towards the next eligible household in the direction indicated by the top of the bottle. To prevent contamination between households in the intervention and control arms, we will ensure a minimum 100m buffer between study households.

### Baseline survey

The study team will administer a baseline survey to record household construction materials, household census, assets, income, animal ownership, WASH infrastructure and practices, demographics, maternal depression, and the illness and activities of a non-index child aged <3 years (index children will be in utero) (Table 2). Surveys will include questions about satisfaction with housing and floor quality. Field staff will visually assess floors for cracks, damage, cleanliness, and the presence of animal feces, food scraps, or trash. Field staff will record participant names and cell phone numbers to assist with tracking should participants relocate during the follow-up period. Field staff will receive standardized training on survey administration and sample collection.

**Table 2:** Summary of variables measured in surveys.

|  |  | Base-<br>line | Birth | 3<br>mo. | 6<br>mo. | 12<br>mo. | 18<br>mo. | 24<br>mo. |
| --- | --- | --- | --- | --- | --- | --- | --- | --- |
| Household characteristics | Household construction materials | X |  |  | X | X | X | X |
|  | Household census | X |  |  | X | X | X | X |
|  | Household assets | X |  |  | X | X | X | X |
|  | Parental education and employment | X |  |  | X |  |  | X |
|  | WASH infrastructure & practices | X |  |  | X | X | X | X |
|  | Animal ownership & husbandry | X |  |  | X | X | X | X |
|  | Animal presence in the home | X |  |  | X | X | X | X |
|  | Human and animal feces in the home | X |  |  | X | X | X | X |
| Perinatal outcomes | Prenatal and birth outcomes |  | X |  |  |  |  |  |
| Anthropometry | Length, weight, head circumference | X | X | X | X | X | X | X |
| Illness symptoms | Child illness symptoms (e.g., diarrhea) | X |  |  | X | X | X | X |
| Flooring | Floor durability assessment | X |  |  | X | X | X | X |
|  | Floor hygiene and maintenance | X |  |  | X | X | X | X |
| Child activities | Child motor skills |  |  |  | X | X | X | X |
|  | Child cognitive development |  |  |  |  | X |  | X |
|  | Child feeding practices |  |  |  | X | X | X | X |
|  | Child play locations |  |  |  | X | X | X | X |
| Maternal outcomes | Quality of life | X |  | X |  | X |  | X |
|  | Satisfaction with housing | X |  |  | X | X | X | X |
|  | Perceived Stress Scale | X |  | X |  | X |  |  |
|  | Edinburgh Postnatal Depression Scale | X |  | X |  | X |  |  |
|  | Maternal bandwidth |  |  |  | X |  | X |  |
|  | Maternal time use |  |  | X |  | X |  |  |
| Flooding status | Household flooding status | X |  |  | X | X | X | X |
| Water, sanitation, handwashing | Water storage, latrine access, child feces management, handwashing | X |  |  | X | X | X | X |
| Vaccination | Vaccination status of index child |  |  | X | X | X | X | X |
| Deworming and antimicrobials | Deworming status of household members | X |  |  | X | X | X | X |
|  | Antimicrobial use of mother and index child | X |  |  | X | X | X | X |
|  | Antimicrobial use in domestic animals | X |  |  | X | X | X | X |
| Food consumption | Animal milk consumption practices |  |  |  | X | X | X |  |
| Environmental sampling | Environmental sample collection |  |  |  | X | X | X | X |

### Randomisation

The trial will use household-level randomisation stratified by geographic block. This design will ensure that intervention and control groups are evenly distributed across the study area and support baseline balance of participant characteristics between arms. Using a block randomised design is also expected to increase study efficiency by accounting for strong geographic clustering of enteric infection (Arnold et al. 2024). As households are enrolled, they will be formed into geographic blocks of 10 spatially contiguous households. There will be no spatial overlap in geographic blocks. Using R statistical software, a Stanford investigator not involved in the data collection will randomly assign households within each block to control and intervention arms in a 1:1 ratio. A random subsample of 50 intervention households stratified by block will be assigned to receive a green concrete floor. Due to the nature of the intervention, it will not be possible to blind participants to a randomised study arm. Laboratory staff and data analysts will be blinded during primary and secondary outcome analyses of intervention effects.

### Intervention

A local non-governmental organization that is not involved in the research will install concrete floors in 400 intervention households before the birth cohort is born. The control group will receive no interventions. The construction team will remove the top layer of the floor and then create a compacted soil foundation. To increase durability during floods and heavy rainfall, the minimum floor height will be 18 inches. They will retain or change the soil foundation of the home according to the household’s preference. The soil foundation will then be covered with a sand layer followed by a layer of bricks and then cement, followed by sealant. The installation process takes approximately 5 days, and then the floor must cure for approximately 7 days. The entire process takes approximately 14 days, and during this time household members will temporarily relocate, and they will return when curing is complete. If it has rained recently, the field team will wait at least 2 days after it stops raining before installing the concrete floor.

In a subsample of the treatment arm (N=50), we will use an alternative cement mix with fly ash that is likely produced with fewer greenhouse gas emissions than ordinary portland cement. 20% of ordinary portland cement will be replaced with fly ash, and commercially produced “green” blocks will be used instead of traditional bricks as supply availability permits. Green blocks will be made of a mixture of cement, fly ash and recycled aggregates.

### Participant retention

To minimize attrition in the control arm, we will compensate mothers from control households with 500 Bangladeshi Taka ($4.27 USD) after completion of each follow-up survey at 3, 6, 12, 18 and 24 months.

### Follow-up measurements

Field staff will conduct follow-up surveys in all intervention and control households at 3, 6, 12, 18 and 24 months (Table 2). They will conduct household surveys, blood spot and stool collection in the full study sample (N=800). The following potential mediators and outcomes will be measured in subsamples: maternal bandwidth, time use and child cognitive development (N=400), environmental samples (N=400 for STH by qPCR, N=110 for STH larvation detected using microscopy, N=110 for *E. coli*, N=110 for extended-spectrum beta-lactamase producing *E. coli*), video observations (N=60) (Figure 2). When selecting subsamples of households for additional measurements, households will be randomly selected stratified by geographic block and study arm. Additional subsamples will be nested within the largest group. The same subsamples will be used across follow-up rounds for those with repeated measures. If any of the households included in the subsamples are lost to follow-up at a particular measurement, they will be replaced with another block-stratified randomly sampled household from the same study arm.

### Birth survey

Field staff will ask study participants to call them once a mother begins labor and when the index child is born. A survey will be conducted as early as possible after birth (up to four weeks from birth) to record obstetric and birth outcomes. If the mother is temporarily residing away from the study household, field staff will travel to the mother’s location if it is within a study upazila. The birth survey includes questions about antenatal care utilization, delivery location, delivery mode, obstetric complications, gestational age, and neonatal mortality. Field staff will measure weight, length, and head circumference.

### Household surveys

At each follow-up, the field team will administer a survey to mothers of the birth cohort to assess any changes to housing materials, household membership, and household assets (Table 2). Follow-up surveys will collect the same measurements as baseline as well as maternal bandwidth, maternal time use, child motor and cognitive development, child anthropometry and child vaccination status. Additionally, mothers will be asked to report index child illness symptoms and daily activities for the index child.

### Stool collection

At 6, 12, 18, and 24 month follow-ups, field staff will provide caregivers with a sterile container to collect stool from the index child. They will return to pick up the collected stool the following morning and will characterize stool consistency using the modified Bristol Stool Form Scale for children (Lane et al. 2011). Field staff will aliquot 0.5g of stool with 1 mL of Zymo DNA/RNA Shield (Zymo Research, Irvine, California) at the household for stool specimens that will be used for metagenomic sequencing. The samples will be transported on ice to the icddr,b laboratory in Dhaka while maintaining a temperature from 4-10°C. For qPCR analyses, 0.5g stool will be aliquoted with 1 mL 100% ethanol and stored at -80°C prior to qPCR analysis. For antimicrobial resistance analyses, stool will be stored at 4°C with no preservatives prior to analysis within 24 hours of specimen collection. After the final round of stool collection at age 24 months, all household members will be offered deworming medication.

### Blood collection

At 6, 12, 18, and 24 month follow-ups, field staff will collect dried blood spots from the index child. After blood spots are dried for 24 hours, they will be transported to the icddr,b laboratory in Dhaka and archived at -80°C for analyses in future studies.

### Environmental sample collection

At 6, 12, 18, and 24 month follow-ups, field staff will collect dust from floors in a subset of households (N=400). In a smaller subset (N=110), the staff will also collect a floor swab and child hand rinse from index children. To collect dust from floors, staff will use a 0.5 x 0.5 m sterilized metal stencil to mark up to 8 adjacent floor areas (total 2 m^2^), starting next to the head of the bed where the index child usually sleeps. They will sweep the area within the stencil with a sterilized brush once vertically and then once horizontally. They will collect the resulting dust using a sterile scoop and place it in a sterile Whirlpak bag. Staff will then mark an adjacent 0.5 x 0.5 m floor area and swab the area within the stencil with a sterile pre-hydrated sponge once vertically and then once horizontally and place the sponge in a sterile Whirlpak bag. To collect index child hand rinses, staff will ask the caregiver to place child’s hands, one at a time, into a sterile Whirlpak bag pre-filled with 250 mL of sterile water. They will massage each hand for 15 seconds from outside the bag, then shake the Whirlpak bag for 15 seconds with the hand submerged.

At the 6 month follow-up only, field staff will collect additional samples in a subset (N=110). This will provide an early assessment of intervention effectiveness on a wider range of matrices as well as detect additional sources of contamination in the domestic environment. The additional sample types include: drinking water (for index child or other children <5 years), prepared food (for index child or other children <5 years), floor soil (in control arm only), courtyard soil, and cow and chicken feces from animals that stay in the compound. To collect drinking water, field staff will ask the caregiver to bring them a glass of water the same way they would give it to the index child, or to another child <5 years if the index child is not yet drinking water. Staff will pour approximately 250 mL of water from the glass into a sterile Whirlpak bag. To collect prepared food, staff will ask the caregiver to bring them a small portion of food the same way they would give it to the index child, or to another child <5 years if the index child is not yet eating food. Staff will use a 50 mL sterile tube with a sterile spoon to collect approximately 50 g of food. To collect floor soil, field staff will use a sterilized metal stencil to mark an additional 0.5 m x 0.5 m floor area near the child’s sleeping area as described above. They will scrape a 5 mL sterile tube along a horizontal line within the stencil to collect approximately 5 g of sample for sequencing. They will then use a 50 mL sterile tube with a sterile spoon to scrape the remaining area within the stencil once vertically and once horizontally; they will repeat these steps until the tube is full to obtain approximately 50 g of sample to determine moisture content and soil type. To collect courtyard soil, field staff will use a sterilized metal stencil to mark a 0.3 m x 0.3 m area at the entrance to the household where the index child lives and follow the same procedure as floor soil collection. To collect cow and chicken feces, field staff will ask the caregiver to identify feces from recent defecation by their animals, then use a 50 mL sterile tube with a sterile spoon to collect the fecal sample. Zymo DNA/RNA Shield (Zymo Research, Irvine, California) will be added immediately after collection to sample aliquots that will be used for metagenomic sequencing. All samples will be transported on ice to the icddr,b laboratory in Dhaka. Samples that will be used for molecular analyses will immediately be stored at -80°C until DNA extraction. Samples that will be used for culture-based analyses will be stored at 4°C overnight prior to analysis by the next day.

### Laboratory assays

We will test for STH in stool samples and household floor dust samples using quantitative polymerase chain reaction (qPCR) using previously published sequences for the following species which had prevalence >5% in our prior research in rural Bangladesh: *Ascaris lumbricoides(Pilotte et al. 2019)*, *Trichuris trichiura*, and *Necator americanus(Pilotte et al. 2016)*. DNA will be extracted from stool samples using the QIAamp Fast DNA Stool Mini Kit (51604, QIAGEN GmbH, Hilden, Germany). Extractions will occur following the manufacturer’s suggested protocol with the following changes: (1) Rather than vortexing, homogenization will occur using the Mini-BeadBeater-24 (BioSpec Products). (2) Immediately following homogenization, 1µL of a 100pg/µL stock of an internal recovery and amplification control (IAC) plasmid (Deer, Lampel, and González-Escalona 2010) will be added to each sample. DNA will be extracted from floor dust samples using the DNeasy PowerSoil Pro kit following the manufacturer’s suggested protocol, again with the addition of IAC plasmid. Following DNA extraction, all samples will be tested for the presence of IAC. Amplification of plasmid target will indicate successful recovery of DNA during the extraction procedure and will demonstrate recovered DNA as amplifiable. DNA extracts that fail to produce an IAC result that falls within 3 SD of the mean of all sample IAC results will be eliminated, and the parent sample will undergo re-extraction. Testing for the presence/absence of IAC will occur in accordance with previously published protocols (Papaiakovou et al. 2018). The presence/absence of STH targets will be determined via the use of previously described small volume, multiparallel qPCR assays. Testing will occur in accordance with published protocols and cycling procedures [60, 61]. Single-well reactions will be used to test all DNA extraction products for the presence of each pathogen of interest, and a qPCR result will be deemed “positive” if testing produces an amplification curve with a Ct value <40.

In a random subsample (N=110) stratified by geographic block, we will determine whether STH ova are larvated using our previously validated method of incubation followed by microscopy since larvated ova can transmit infections to humans, whereas non-larvated ova cannot (Steinbaum et al. 2017).

In the same subsample (N=110), we will use IDEXX Quanti-Tray/2000 with Colilert-24 (IDEXX Laboratories, Westbrooke, Maine) to enumerate the most probable number (MPN) of *E. coli* in floor swab and child hand rinse samples. Swabs will be eluted by adding 100 mL of sterile water to the Whirlpak bag containing the sponge, massaging the swab from outside the bag for 15 seconds, then swirling the Whirlpak bag for 15 seconds. This step will be repeated three times to generate 300 mL of eluate (Harris et al. 2016). 5 mL of the eluate will then be diluted with 95 mL of sterile water to generate a 100 mL aliquot for IDEXX testing. 50 mL of child hand rinse water will be diluted with 50 mL of sterile water to generate a 100 mL aliquot for IDEXX testing. For floor swab samples, a second 100 mL aliquot will be processed after adding 80 μL of filter-sterilized 5 mg/mL cefotaxime solution (final concentration of 4 μg/mL in the 100 mL aliquot) to enumerate the MPN of cefotaxime-resistant *E. coli* (Hornsby et al. 2023).

In the same subsample (N=110), we will also detect and enumerate extended-spectrum beta-lactamase producing *E. coli* (ESBL-E) in the following sample types: child stool, cow and chicken feces from the same compound, floor swab, courtyard soil, child hand rinse, drinking water, and prepared food. We will use CHROMagarTM ESBL media to isolate and identify ESBL-E (Hossain et al. 2021). Selected colonies from each plate will be analyzed by qPCR for major ESBL genes (CTX-M, SHV, TEM, OXA) following previously published protocols (Mahmud et al. 2022). Isolates from previous studies will be used as positive controls. For selected isolates, antibiotic susceptibility will be determined using the Kirby-Bauer disk diffusion method following guidelines of the Clinical and Laboratory Standards Institute (CLSI) and European Committee on Antimicrobial Susceptibility Testing (EUCAST) (“CLSI Publishes M100—Performance Standards for Antimicrobial Susceptibility Testing, 32nd Edition,” n.d., “Breakpoint Tables for Interpretation of MICs and Zone Diameters” 2021).

In the same subsample (N=110), we will perform metagenomic sequencing of child stool, floor soil, cow feces and chicken feces samples from each follow-up round. Floor soil will only be collected from control arm households (N=55) because we expect that the quantity of soil needed for DNA extraction (10 g) will not be available on concrete floors. We will combine 10 fecal samples of each type and 5 floor soil samples to generate 11 composite samples of each type. We will extract DNA from each composite sample using DNeasy PowerMax Soil Kits for soil samples and Qiagen QIAamp PowerFecal Pro DNA Kits for stool samples. Metagenomic shotgun sequencing will be performed using the Illumina platform. We will use Kraken2 (Wood, Lu, and Langmead 2019) to perform taxonomic classification of reads to identify pathogens and bacterial antimicrobial resistance genes in each sample.

### Maternal measures

We will measure maternal stress, depression, and time use at child ages 3 and 12 months and maternal bandwidth at child ages 6 and 18 months. This sequencing will minimize participant fatigue by reducing the number of measures collected per round and will ensure that mediators are measured prior to each child cognitive measurement at 12 and 24 months.

Maternal bandwidth measures will include visual-spatial working memory, executive function, inhibitory control, and cognitive flexibility and attention shifting. To measure visual-spatial working memory, we will use an analog version of the Corsi Block Span Task (Berch, Krikorian, and Huha 1998) since mothers may be unfamiliar with computers or tablets. The analog version is a customized set of blocks located in fixed positions on a board. An enumerator will tap on a sequence of blocks and mothers will be asked to recall the sequence by tapping the blocks. Sequence length will increase in each round, with a maximum sequence length of nine blocks for the forwards Corsi task and eight blocks for the backwards Corsi task, where the mother recalls the sequence in the reverse order. Field staff will record the maximum sequence length correctly recalled by the mother.

We will measure perception, abstraction capacity, analogical reasoning and metacognitive executive function except emotion using Raven’s Progressive Matrices (Diamond 2013; John and Raven 2003). Each booklet will contain 48 patterns with a missing tile inside. Mothers will have 45 minutes to choose the missing tile that best completes each of the 48 patterns. At the end of the task, the total raw score will be converted to an Ability Score and Standard Score.

We will also measure cognitive inhibition, which refers to the ability to suppress the processing of stimuli that are irrelevant to the intent of the task and inhibit a prepotent response. We will use an adapted version of The Hearts and Flowers task (Davidson et al. 2006) using images of cats and dogs, which are recognizable to the study population. This task takes into account both accuracy and speed and captures individual differences in inhibition and cognitive flexibility (Davidson et al. 2006; Obradović et al. 2017). This task has three rounds, each with 20 trials. In the first round, dogs will appear randomly on either the right or left side of a page, and respondents (mothers) will be asked to respond to one rule: to place their hand over an arrow on the same side of page as a dog image (congruent task). In the second round, cats will appear on either side of the page, and mothers will be asked to place their hand on an arrow on the side opposite to where the cat appears (incongruent task). In the third round, dogs and cats will appear intermixed (mixed task), and mothers will be asked to respond accordingly. Usually, no executive demands are required in the Dog task, thus it is used as a control task to compare performance on the Cat and mixed tasks. The percentage of correct responses and response time will be recorded for each respondent.

To measure cognitive flexibility and attention shifting, we will use Dimensional Change Card Sort (DCCS) test presented as stimuli on flip charts instead of a screen (von Suchodoletz, Slot, and Shroff 2017). Two target pictures will be presented that will vary along two dimensions (e.g., shape and color). The participants will match the picture at the top of the displayed page with two pictures with varied dimensions (e.g., shape and color) at the bottom. To match the picture according to the requested dimension spoken by the tester, participants will place their hand on an arrow at the side of the correct corresponding picture. Following practice trials with color-matching and shape-matching, 29 mixed trials will be offered. We will record the percentage of correct responses and response time for each participant.

To measure maternal discretionary time, we will use a hybrid time diary adapted to our setting in Bangladesh (Erica M. Field, Rohini Pande, Natalia Rigol, Simone G. Schaner, Elena M. Stacy, Charity M. Troyer Moore 2022). Enumerators will invite mothers to summarize the prior day’s activities chronologically and allocate hours by placing tokens next to photos of 9 common activities. This approach does not require mothers to perform calculations or be literate. We will calculate total time spent on each activity to determine discretionary time.

### Child development assessment

The Ages and Stages Questionnaire Inventory (ASQ:I) will be administered at child ages 12 and 24 months to evaluate problem solving, communication, fine motor, gross motor, and personal social development in index children. The ASQ:I is a continuous sliding scale with questions for each domain and starts with the child’s chronological age followed by establishing a basal and ceiling levels of a child’s performance. Although it is a tool based on parents’ report, in Bangladesh, the ASQ:I was adapted by researchers to include a subset of items that are administered through direct assessment with locally available materials that the parents may not observe. Administration of the ASQ:I requires adequate training and around 45 minutes to administer. The adapted version of this instrument has been validated in Bangladesh (Pitchik et al. 2023). The Family Care Indicator (FCI) instrument will be used to measure the amount of stimulation the child receives at home at 12 and 24 months. This instrument measures stimulating activities, the availability of a variety of play materials, caregiver responsiveness and evaluates the child’s surrounding environment. This instrument has been validated in Bangladesh (Hamadani et al. 2010).

### Quality assurance for psychosocial measures

Child development and maternal bandwidth data will be collected by trained physicians and psychologists from icddr,b. All measurements will begin after pretesting in the community on non-study mother-child dyads (N=40), once an inter-observer agreement value of 90% is achieved between testers and trainers. Refresher training will take place every 6 months or when correlation coefficients for tester-supervisor pairs fall below 0.85. In a random sample of 5–10% of tests, testers’ evaluations will be compared to evaluations completed by trained psychologists.

### Video recording and analysis

We will record 6-hour video observations in a random subsample of 60 children (30 children per arm). Recording start times will vary from 7 AM to 12 PM between households to capture a wider range of activities and will be conducted in the household, courtyard, or near the courtyard. Observations will be paused during breastfeeding, bathing, and sleeping times. Videos will be coded to measure the duration, time, and location of relevant child activities, such as the soil contact and ingestion and animal contact, using LiveTrak software (Julian et al. 2018).

### Data management

Data will be collected on tablets using customized CommCare software, which facilitates longitudinal data collection. Data will be securely stored on CommCare servers and at icddr,b, with access limited to the study PI and the Data Manager. Paper records will be digitized and destroyed, and personal identifiers will be removed. Digitized materials will be stored indefinitely, with identifiable information password-protected and encrypted. Hard copies will be stored in locked cabinets. To ensure data quality, supervisors will observe interviews, conduct re-interviews, cross-check data, and provide feedback to enumerators. Field staff will maintain a field tracking form to track daily data collection activities. Stata and R scripts will regularly be used by the data manager, investigators, and statisticians to identify inconsistencies and errors, and errors will be resolved prior to analysis.

### Statistical analysis

To assess randomisation, we will compare the means and percentages of baseline characteristics between arms including maternal age, maternal education level, paternal education, paternal employment, housing wall and roof materials, household assets, acres of homestead land owned, household WASH infrastructure and access, open defecation practices, and deworming in the prior 6 months, animal ownership, and animal cohabitation.

The primary analysis will compare prevalence of any STH between study arms using an indicator for whether a child tested positive at the 6, 12, 18, or 24 month follow-up rounds. We will estimate the unadjusted prevalence ratio comparing intervention to control using generalized estimating equations (Zeger, Liang, and Albert 1988) with a binomial family and log link and an exchangeable correlation matrix to account for repeated measures within the same child. If log-binomial models do not converge, we will use modified Poisson regression with robust standard errors (Zou 2004). Secondary analyses will include indicators for follow-up rounds in order to estimate the prevalence ratio at each follow-up to assess whether effects vary over different child ages. All models will condition on geographic block and estimate robust sandwich standard errors (Freedman 2006) to account for the matched design (McNutt et al. 2003; Kernan et al. 1999). To assess whether the intervention effect is sustained over 2 years, we will also fit a model with an interaction term between treatment and follow-up time.

We will also compare secondary outcomes (Table 1) between study arms. For repeated measures outcomes, we will use generalized estimating equations (Zeger, Liang, and Albert 1988) with an exchangeable correlation matrix; we will estimate mean differences for continuous outcomes using an identity link and Gaussian family and prevalence ratios for binary outcomes using a log link and binomial family (McNutt et al. 2003). For measurements analyzed at a single time point, we will fit generalized linear models (McNutt et al. 2003) with a Gaussian family and identity link for continuous outcomes and a binomial family and log link for binary outcomes. If log-binomial models do not converge, we will fit models with a Poisson and log link (McNutt et al. 2003; Zou 2004).

Primary analyses will be unadjusted for covariates. A secondary analysis will adjust for baseline covariates to reduce bias associated with any chance imbalances between study arms at baseline and possibly increase efficiency (Pocock et al. 2002). We will pre-specify a potential covariate adjustment set of baseline variables and only adjust for covariates among this set that are associated with each outcome (likelihood ratio test p-value < 0.2).

Analyses will be intention-to-treat. The following households will be considered non-compliant: 1) control households that install concrete floors that cover at least one entire room inside the mother’s household; 2) households that cover at least one room’s floor with a cement finish; 3) intervention households that remove any part of the concrete floor installed as part of the trial; 4) intervention households in which a substantial portion of the concrete floor is damaged. If non-compliance exceeds 10% in either arm, we will perform a per-protocol analysis as well. We will not adjust for multiple comparisons because the number of outcomes is relatively modest, and the analysis will follow a pre-specified analysis plan.

We will compare attrition rates at each follow-up and across follow-ups between randomised arms and compare baseline characteristics of those lost to follow-up to those who complete the study. If there is evidence of systematic differences in attrition between arms and attrition exceeds 20%, we will explore whether study drop out resulted in selection bias using inverse probability of censoring weights with flexible machine-learning based estimation (Mark J. van der Laan 2011).

For STH measured in floor dust samples, the primary analysis will impute STH prevalence as 0 for floors that do not contain enough dust for nucleic acid extraction (0.1 g). The imputation assumes that floors with no dust do not harbor STH eggs. Our pilot data indicate that concrete floors in 40% of households may yield insufficient dust for nucleic acid extraction. We will conduct a secondary analysis by comparing STH outcomes in floor dust between intervention and control groups only using data from households where sufficient dust was obtained. We expect this analysis to yield a conservative estimate of intervention effectiveness because it will exclude intervention households with dust-free floors which are less likely to harbor STH eggs.

We will explore potential effect modification by the following baseline variables: household wealth, household WASH infrastructure, presence of animals inside the household, primary play location of children <2 years, index child sex, household distance to nearest permanent water body (Pekel et al. 2016), and ordinary portland cement vs. green cement. We will also assess effect modification by monsoon vs. dry season using a data-based definition and precipitation, temperature, and humidity levels (Nguyen et al. 2024) using weather station data from the Government of Bangladesh Meteorological Department. We will test for effect modification on the relative scale by including interaction terms between these variables and treatment group indicator variables in regression models. We will estimate additive scale interactions from log-linear models using the relative excess risk due to interaction (RERI) (Knol et al. 2011). To test for effect modification, we will estimate p-values on the interaction terms for relative scale interaction in log-linear models and for additive scale interaction RERIs (Knol et al. 2011; Assmann et al. 1996).

To assess whether any post-treatment variables mediate intervention effects, we will perform causal mediation analyses (Tingley et al. 2014) for the following potential mediators: deworming or antimicrobial consumption of birth cohort or other household members in the past 6 months, and floor coverings. Interim reductions in STH could result in reduced deworming use, which in turn, further reduces STH at the final follow-up measurement. Additionally, installation of concrete may alter the type and frequency of floor coverings used, which may influence health outcomes. We will also use causal mediation analysis to assess whether health and child development outcomes are mediated through environmental vs. maternal pathways. For the environmental pathway, mediators will include contamination of the home with *E. coli* and STH. For the maternal pathway, mediators will include maternal discretionary time use; maternal bandwidth (measures of executive function, inhibition, and working memory); and maternal quality of life, stress, and depression.

For the ASQi outcome, if child age varies substantially at each follow-up round, we will use age-specific Z-scores for each test within 2-month age bands and will control for age in the analysis. We will create reference distributions for each ASQi domain (communication, fine motor, gross motor, problem solving, and personal social) and the overall global scale. The sum of age-specific raw scores from the control group will be standardized with a mean of 0 and a SD of 1 for each 2-month age band. For the intervention arm, we will calculate standardized Z-scores using the reference distribution from the control arm in each age band. We will summarize the distribution of continuous outcomes (e.g., ASQi Z-scores) using Gaussian kernel density smoothers. We will estimate mean differences in ASQi scores or Z-scores between study arms using generalized linear models that condition on geographic block. To compare rates of attainment for each growth milestone, we will estimate hazard ratios using a semiparametric generalized additive model with a complementary log-log link and baseline hazard fit with a monotonic cubic spline.

### Cost-effectiveness analysis

We will conduct cost-effectiveness to examine both the costs and health outcomes of the concrete floor intervention. We will collect data on floor installation costs from a program perspective, separating material and labor costs, since in the future households may install floors themselves. Using an ingredients-based approach, we will record costs for individual line items within categories including capital costs, materials, transportation, personnel, and operations. We will then adapt our previously published natural history and cost-effectiveness models of childhood helminth infections and enteric fever to project the health impacts and cost-effectiveness of floor installation (Lo et al. 2016, 2018; Ryckman et al. 2021; Weyant et al. 2024). We will populate the model with outcome rates from the control and intervention arms of this trial, simulate age-specific risk over a ten-year period, and project DALYs for each health outcome for households with concrete floors and soil floors.

We will calculate DALYs for child STH infections, child diarrhea, child wasting, and maternal depression using published disability weights (Global Burden of Disease Collaborative Network/Institute for Health Metrics and Evaluation, n.d.). While we expect the intervention to influence a broad range of outcomes, disability weights are only available for the subset listed above. We will calculate disability weights for hookworm-associated anemia as the product of the disability weight for anemia due to hookworm disease by the probability that an individual infected with hookworm develops anemia (Bartsch et al. 2016). For each STH infection, we will estimate the probability of mild, moderate, and severe infections based on Ct values from qPCR using data from a prior study of children in rural Bangladesh (Benjamin-Chung et al. 2020). For diarrhea, we will calculate weights assuming the distribution of mild, moderate, and severe diarrhea is the same as found in the MAL-ED study in Bangladesh (Platts-Mills et al. 2015).

We will perform simulations under different scenarios for the durability of intervention effects and intervention coverage, adjusting for inflation. The model will predict costs and health outcomes, deaths, and DALYs averted over a 10-year period; we will obtain 95% credible intervals for uncertainty. We will estimate the incremental cost-effectiveness ratio for concrete vs. soil household floors. Parameter draws from variable distributions will be performed through Latin Hypercube Sampling (Stein 1987). We will perform a probabilistic sensitivity analysis with pre-specified ranges of key parameters (e.g., disability weights, costs, intervention effects) supported by our study findings and the literature (Blower and Dowlatabadi 1994; Basu and Andrews 2013). If we find that the cost of concrete floors made with the fly ash mix differs from that of traditional concrete floors, we will repeat the cost-effectiveness analysis specifically for green concrete floors using the same methods.

## Data Availability

There is no data associated with this trial protocol.

## Ethics and dissemination

The trial protocol was approved by the International Centre for Diarrhoeal Disease Research, Bangladesh (icddr,b) Ethical Review Committee (PR-22069) and the Stanford Institutional Review Board (63990).

The US- and Bangladesh-based PIs will be responsible for safety oversight. PIs will meet twice each year. The study’s data collection phase will end after the final measurements are taken when children are approximately 2 years of age. The PIs will continue to monitor the study population for two weeks after offering deworming to household members at the end of the data collection period in case of adverse events. Interim analyses will only be conducted at the discretion of the PIs to ensure safety of participants and to plan for future data collection and analyses.

The results of the study will be presented at stakeholder workshops in Bangladesh, which will include community members, local, regional, and national stakeholders. In addition, results will be presented on ClinicalTrials.gov, in scientific manuscripts, and at scientific conferences. De-identified study data and replication scripts will be made publicly available online at the time of manuscript publication.

## Author statement

JBC, MR, and AE conceptualised and developed the study and its design. All authors contributed to protocol development. MR, FJ, SH, AY, FT, AE, and JBC wrote the protocol. MR, FJ, SH, AE, and JBC wrote the manuscript. All authors approved the manuscript final draft.

## Funding statement

Research reported in this publication was supported by grants to JBC from the Eunice Kennedy Shriver National Institute of Child Health & Human Development of the National Institutes of Health under Award Number R01HD108196, the Stanford Woods Institute for the Environment (281131), the Rosenkranz Prize, and the McCormick and Gabilan Fellowship; and a seed grant to AE from the North Carolina State University Global One Health Academy. JBC is a Chan Zuckerberg Biohub Investigator. icddr,b acknowledges with gratitude the commitment of Grand Challenges Canada to its research efforts. icddr,b is also grateful to the Governments of Bangladesh and Canada for providing core/unrestricted support.

## Disclaimer

The content of this manuscript is solely the responsibility of the authors and does not necessarily represent the official views of the National Institutes of Health.

## Conflict of interests statement

The authors have no competing interests to declare.

## Notes

### Competing Interest Statement

The authors have declared no competing interest.

### Clinical Trial

NCT05372068

